# Probable causes and risk factors for positive SARS-CoV-2 test in recovered patients: Evidence from Brunei Darussalam

**DOI:** 10.1101/2020.04.30.20086082

**Authors:** Justin Wong, Wee Chian Koh, Riamiza Natalie Momin, Mohammad Fathi Alikhan, Noraskhin Fadillah, Lin Naing

## Abstract

We report findings of a national study in Brunei Darussalam indicating that one in five recovered patients subsequently test positive again for SARS-CoV-2—this risk is nearly three times higher in older patients (age 53 and above) than younger ones (below age 53). Review of clinical and epidemiological records do not support reinfection or reactivation as likely causes of the ‘re-positive’ observation. Instead, prolonged but intermittent viral shedding is the most probable explanation. We discuss the implications of these findings for infection control and clinical practice.

## Introduction

Most criteria for de-isolation of COVID-19 patients recommend two negative RT-PCR tests from respiratory specimens at 24-hour intervals at least 8 – 14 days after symptom onset.^1^ While there are reports of patients who have been discharged and subsequently report positive RT-PCR, no study has attempted to describe the magnitude and significance of this issue at the country level.^2,3,4,5^ This phenomenon which we term ‘re-positive’ could complicate the management of COVID-19 patients.

Up till April 12, 2020, Brunei Darussalam recorded a total of 136 COVID-19 cases of which 106 have been discharged. We follow-up all discharged patients, identify those who are re-positive, describe their clinical and epidemiological outcomes, and analyse the predictors of re-positive status.

## Methods

### Patient diagnosis and management

We confirmed SARS-CoV-2 by RT-PCR assay on nasopharyngeal (NP) specimens if the cycle threshold (Ct) values for Orf1ab was <40. A commercial kit (BGI Genomics) was used.

Newly diagnosed patients are admitted to the National Isolation Centre (NIC) and monitored for at least 14 days from admission until two consecutive negative specimens collected at least 24 hours apart. Discharged patients are required to self-isolate at home, initially for a period of 7 days and later extended to 14 days. They undergo NP specimen collection at day 11 post-discharge. For patients already past day 11, NP specimens were collected at the earliest opportunity. Discharged patients found to be re-positive were readmitted. Bloods, chest X-ray, and antibody testing using the VivaDiag Rapid Test were conducted on readmission.

All close contacts (defined as any person living in the same household, or was within 1 meter in an enclosed space for more than 15 minutes) of the re-positive cases were tested with RT-PCR.

### Empirical analysis

We review all discharged cases that had their post-discharge swab. We calculate the re-positive rate (percentage) by gender, age, clinical severity on first admission, and use of antiretroviral treatment (400 mg lopinavir/100 mg ritonavir) on first admission, and apply log-binomial models to obtain risk ratios. We also compare pre- and post-discharge RT-PCR Ct values of re-positive patients using a paired t-test.

## Results

### Clinical and epidemiological characteristics of re-positive patients

106 patients had a follow-up NP swab taken between 11 - 18 days post discharge. 21 (19.8%) were found to be re-positive (Figure 1). 12 were male and 9 female. The median age was 47.0 years. 20 (95%) of the re-positive patients were asymptomatic, and only one (Patient D) reported symptoms (a mild cough) post-discharge. Routine blood and chest x-ray for all re-positive patients were normal. 14 (67%) of the re-positive patients had both IgM and IgG detected.

**Figure 1.**
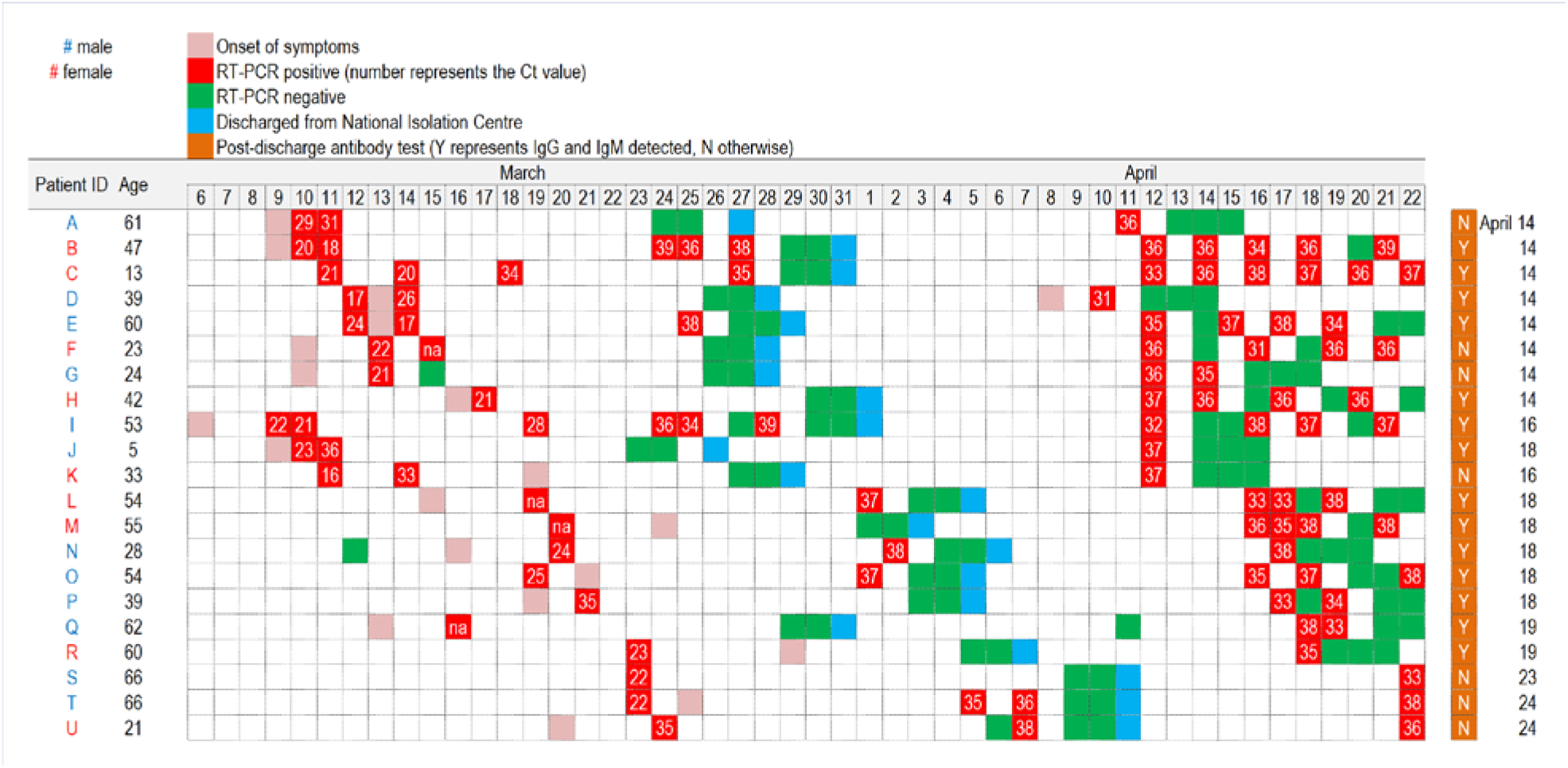
Timeline of re-positive cases following discharge

We used Ct value as a proxy for viral load, with the value inversely related to RNA copy numbers.^6^ The average Ct value of re-positive patients was lower pre-discharge compared to their readmission Ct value. This was statistically significant (p-value = 0.000) (Table 1).

**Table 1.** Comparison of RT-PCR Ct values of re-positive patients pre- and post-discharge

|  | No. of patients | Mean Ct value | (95% CI) | p-value<sup>a</sup> |
| --- | --- | --- | --- | --- |
| Pre-discharge | 19 | 27.78 | (25.28, 30.27) |  |
| Post-discharge | 19 | 35.58 | (34.80, 36.36) |  |
| Difference | 19 | -7.80 | (-10.20, -5.41) | < 0.001 |

Note: Pre-discharge Ct values of two patients unavailable <sup>a</sup> Paired t-test

Contact tracing identified 111 close contacts. All were tested for SARS-CoV-2. One household contact of Patient D tested positive; however, epidemiological investigation concluded that the likely exposure had occurred more than a month earlier.

### Factors associated with re-positive status

We compared re-positive rates of subgroups of four variables in Table 2. The highest re-positive rate was observed in patients aged 53 and above (38.5%), followed by those with moderate to critical conditions (33.3%) and lopinavir/ritonavir treatment (30.3%).

**Table 2.** Comparison between discharged patients who tested positive and those that remained negative

|  | RP<sup>a</sup> | Neg<sup>b</sup> | RP%<sup>c</sup> | Crude RR<sup>d</sup> | (95% CI) | Adj. RR<sup>e</sup> | (95% CI) | p-value |
| --- | --- | --- | --- | --- | --- | --- | --- | --- |
| **Gender** |  |  |  |  |  |  |  |  |
| Male | 12 | 52 | 18.8 | 1.00 | - | 1.00 | - |  |
| Female | 9 | 33 | 21.4 | 1.14 | (0.51, 2.47) | 1.34 | (0.61, 2.71) | 0.423 |
| **Age (years)** |  |  |  |  |  |  |  |  |
| 0 - 52.9 | 11 | 69 | 15.9 | 1.00 | - | 1.00 | - |  |
| 53 and above | 10 | 16 | 38.5 | 2.80 | (1.31, 5.93) | 2.94 | (1.32, 6.32) | 0.005 |
| **Clinical condition** |  |  |  |  |  |  |  |  |
| Asymptomatic | 3 | 26 | 10.3 | 1.00 | - | 1.00 | - |  |
| Mild | 13 | 49 | 21.0 | 2.03 | (0.72, 8.35) | 2.27 | (0.83, 9.13) | 0.165 |
| Moderate, severe, or critical | 5 | 10 | 33.3 | 3.22 | (0.92, 14.11) | 2.44 | (0.73, 10.47) | 0.173 |
| **Lopinavir/ritonavir treatment** |  |  |  |  |  |  |  |  |
| No | 11 | 62 | 15.1 | 1.00 | - | 1.00 | - |  |
| Yes | 10 | 23 | 30.3 | 2.01 | (0.93, 4.32) | 1.52 | (0.56, 3.50) | 0.333 |

<sup>a</sup> RP = Re-positive
<sup>d</sup> Crude risk ratio (Simple log-binomial model)
<sup>b</sup> Neg = Negative
<sup>e</sup> Adjusted risk ratio (Multiple log-binomial model)
<sup>c</sup> Re-positive rate in percent

Multivariable log-binomial model identified age as the only significant risk variable. The best cut-off point was age 53, to use as the dichotomous variable, for modelling re-positive risk (Figure 2).^7^

**Figure 2.**
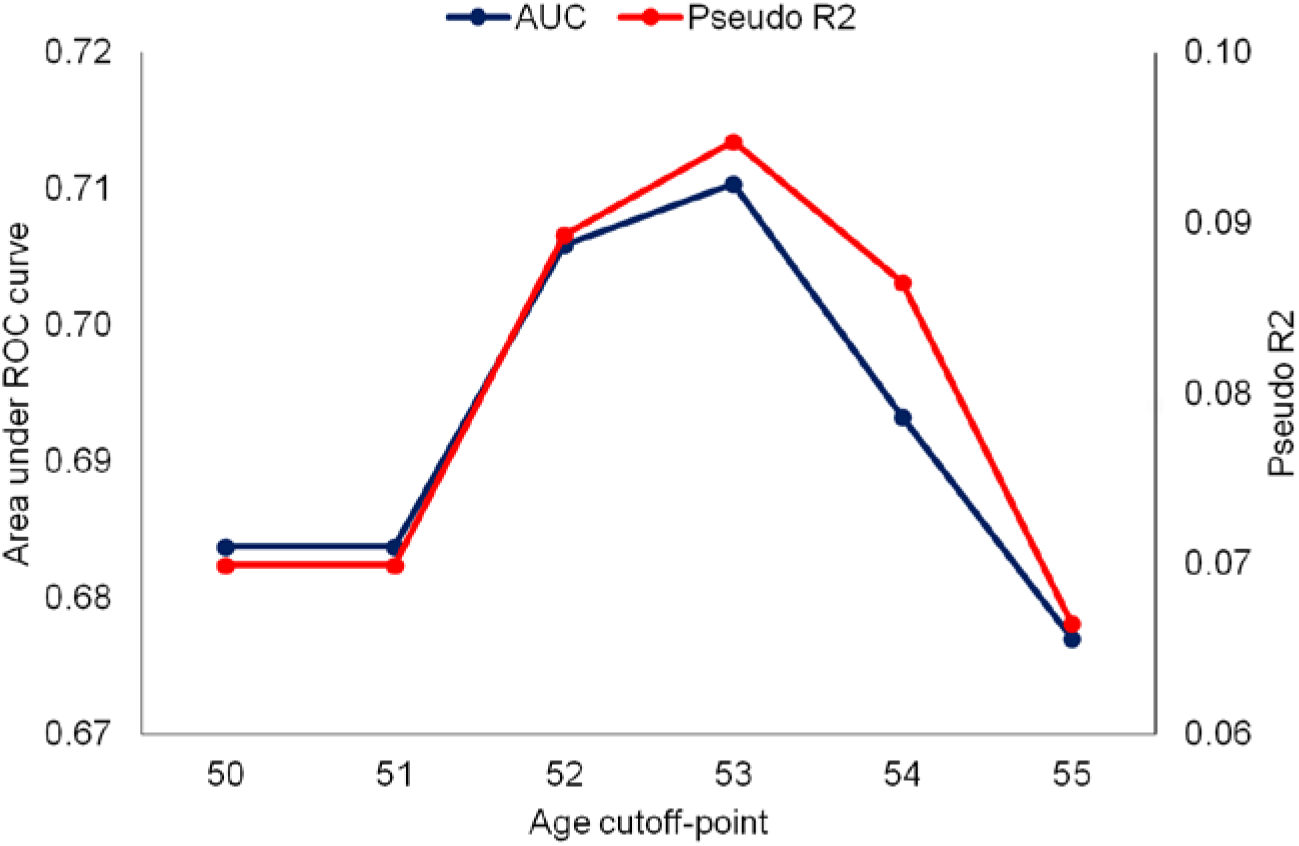
Model fit for re-positive risk by age

Although statistically not significant, the following may well have clinical significance: females had about one-third higher re-positive risk than males, patients with symptoms had at least twice the risk of asymptomatic cases, and those who were given lopinavir/ritonavir had more than 50% higher risk than those not given.

## Discussion

### Key findings

We report a 19.8% re-positive rate. Our study is the first to identify such a high proportion in a cohort, and also the only country-level study that has followed up all discharged cases with post-discharge swab. This could explain the high re-positive rate in Brunei.

There were some observed differences between the two groups (re-positive and consistent negative). The associations with clinical severity and lopinavir/ritonavir treatment were strongly attenuated in the multivariable model suggesting age as the key variable under consideration.

### Sampling and detection of SARS-CoV-2

We cannot exclude test performance or operator deficiencies in specimen collection as a contributing factor to our observed high re-positive rate. NP swabs may be less sensitive for SARS-CoV-2 during the convalescent period and as such could have resulted in false negatives on initial discharge - however the need for two consecutive negatives should mitigate against this.^8^ Also, sampling and detection deficiencies cannot explain the higher risk in older individuals.

### Reinfection, reactivation, residual infection

Some reports suggest reinfection as a possible cause; our findings do not support this.^9^ There was no evidence of infection among close contacts which would have been likely if they were reinfected (as there would have to be an infective source). Moreover, 67% of re-positive patients in our study had antibodies on admission (although this does not necessarily correlate to protection).^10^

20 (95%) patients were asymptomatic on re-detection, and negative RT-PCRs were observed in 16 patients soon (within 1 – 3 days) after they were readmitted, suggesting that reactivation (a phenomenon not observed in other human coronaviruses) is also unlikely.

Our findings support prolonged but intermittent viral shedding as the most plausible explanation. First, we observed oscillation between positivity and negativity, particularly when Ct values were at the detection limit. Second, prolonged viral shedding is observed in SARS-CoV-2 positive individuals up to 37 days after the onset of symptoms among adult patients, consistent with our observations of repeat positive RT-PCR at 27 – 34 days following first diagnosis.^11^ Third, older patients are more likely to have severe disease and to encounter prolonged viral RNA shedding, consistent with the higher re-positive risk among older patients.^12 13^

### Infectivity

Viral RNA shedding of SARS-CoV-2 does not equate with infectivity. While we did not culture the samples in our study population, other attempts at live virus isolation found that no isolates were obtained after day 8.^14^ Our epidemiological findings support the virological observations—of the 111 close contacts tested, none were found to be positive as a result of exposure to a re-positive patient.

### Implications for clinical practice

We observed that the re-positive rate is higher than commonly reported, with increased risk in older age groups. Our findings support prolonged but intermittent viral shedding as the probable cause for this phenomenon. While we did not observe infectivity potential in these patients, given the high re-positive rate observed, it would be dangerous to exclude this possibility entirely. Based on our findings, we suggest that patients should be isolated for an extended period of time even after discharge (until at least 28 days from first diagnosis). Where capacity permits, they should be tested for SARS-CoV-2 RNA prior to full de-isolation, particularly those with a higher re-positive risk such as in older individuals.

## Data Availability

The data of the discharged patients are not publicly available. A summary is presented in Table 1, and a timeline of the RT PCR test results of the re-positive patients are in Figure 1.

